# Brain-Only Versus GI-Only Synucleinopathy: A Comprehensive Autopsy Study With Both IHC and SAA

**DOI:** 10.64898/2026.03.18.26348355

**Authors:** Christina D. Orru, Thomas G. Beach, Charles H. Adler, Holly A. Shill, Erika Driver-Dunckley, Shyamal H. Mehta, Alireza Atri, Ileana Lorenzini, Sanaria H. Qiji, Anthony J. Intorcia, Andrew G. Hughson, Bradley R. Groveman, Samantha King, Parvez Alam, Sabiha Parveen, Sarah Vascellari, Byron Caughey, Geidy E. Serrano

## Abstract

Braak and others have proposed that Lewy body pathology (LBP) in Parkinson’s disease (PD) may arise not only in the brain but alternatively from an initial site in the gastrointestinal (GI) tract with subsequent passage to the central nervous system CNS through the vagus nerve or other routes. We tested this hypothesis by using both immunohistochemistry (IHC) and RT QuIC a form of alpha synuclein seed amplification assay (SAA) to detect alpha synuclein LBP in samples from selected brain regions and 10 GI tract sites taken from autopsies of 50 PD subjects and 128 elderly subjects without parkinsonism or dementia including 34 with IHC identified CNS incidental Lewy body disease (ILBD) and 94 with no Lewy body IHC pathology detected (NLB). A positive SAA or IHC result was restricted to the GI tract in only 2 subjects while LBP by either SAA or IHC was restricted to the brain in 11 subjects. To fairly compare GI-only with brain-only synucleinopathy, however, we would have to do SAA on brain samples from all ILBD and NLB cases in at least 4 critical brain regions: olfactory bulb, medulla, pons, and amygdala. Further SAA of brain regions is estimated, based on the proportional results to date, to potentially identify 21 additional brain-only LBP subjects, for a total of 32, if it were done on all of the NLB subjects. From this brain-only LBP is estimated to be 16 times more common than GI-only LBP. To assess the clinical impact of SAA-positive GI sites we found that the number of positive sites per subject is significantly correlated with UPDRS motor score and SCOPA-AUT GI related scores including those for salivation, straining, constipation, and bowel movement.

## INTRODUCTION

In the first half of the 20^th^ century, not long after Fritz Lewy, Gonzalo Lafora and Konstantin Tretiakoff first described Lewy bodies (1–4) in the brains of people with Parkinson’s disease (PD), others, including Herzog, Wohlwill, and Hechst and Nussbaum, and later den Hartog Jager and Bethlem (5), reported them also in the peripheral nervous system (PNS), within ganglion cells of the sympathetic vertebral chain. Decades later, Qualman et al (6), Lipkin et al (7) and Wakabayashi and colleagues (8) reported Lewy bodies in ganglia of the GI tract (9). After the discovery that α-synuclein was the major constituent of Lewy bodies, sensitive immunohistochemical (IHC) methods demonstrated that the distribution of “α-synucleinopathy” extends widely throughout the body (10–12). This revelation then instigated a cascade of research aimed at using PNS α-synuclein pathology as a biopsy biomarker as well as raising the novel question as to whether or not PD begins in the brain or in the body (13–19). The stimulus for the latter alternative, which has been called the “body-first” hypothesis, initially came from clinical PD studies that found a wide range of non-motor signs and symptoms that accompany or may even precede the motor signs (20–24).

Many of these non-motor accompaniments are related to gastrointestinal (GI) dysfunction and therefore much attention has been focused on the stomach as the major “first stop” along the alimentary tract and hence the most likely place for the initiation of synucleinopathy, perhaps by absorption of toxins or through microbial or inflammatory stimuli, followed by transmission to the brain through the vagus nerve (19). Supporting this is the repeatedly-confirmed finding of a rostrocaudal GI gradient of synucleinopathy (8,11,25,26), which may itself be related to the known distribution of vagal GI innervation (27). A stomach-vagal-brain connection has been invoked to explain epidemiological reports that subjects with prior vagotomy may have a lower prevalence of PD, although this has been disputed (28–35). Similarly, although the normal appendix may have a high concentration of α-synuclein (15), there is conflicting epidemiological evidence of a possible protective effect of appendectomy (36–46). A stomach “inoculation” site has been experimentally tested in multiple animal models, and a consensus has emerged that bidirectional spread is possible, both upwards from the stomach and downwards from the brainstem (47–49).

Based on theoretical clinical progression models, Borghammer and colleagues (50–54) proposed the existence of two subtypes of PD, one that is “brain-first” and one that is “body-first”. A critical weakness of the body-first hypothesis, however, has been that published autopsy studies including our own initial multi-organ study (11) and our more recent study focused on stomach and vagus nerve (55) have indicated that peripheral synuclein pathology very rarely exists in the absence of brain synuclein pathology. The only autopsy support cited by the Borghammer group (51) comes from a few Japanese studies (56–58) finding isolated synuclein pathology in the sympathetic ganglia of elderly subjects but still lacking evidence of body-only GI synuclein pathology. The most convincing of these autopsy reports is that of Tanei et al (56), which included 178 autopsy cases with representation of both CNS and PNS sites. The latter included sections of adrenal, heart, sympathetic ganglia, esophagus and skin. Only 9 cases were exclusively positive in the PNS (9/178 = 5%) while 121/78 (68%) had both CNS and PNS pathology, and 48/178 (27%) were only CNS-positive. All of the PNS-only cases were only positive in sympathetic ganglia or cardiac innervation. While this supports the possible rare existence of body-first cases, isolated synuclein pathology was 6-fold more common in the CNS, and the first PNS location would seem to be within the sympathetic nervous system, not the GI tract as has been extensively proposed. It seems logical that catecholaminergic PNS neurons would have a selective inherent vulnerability to synucleinopathy, similarly to catecholaminergic CNS neurons, not necessarily invoking an exogenous pathogen.

A remaining conundrum for the body-first hypothesis would be how to explain the spread of synuclein pathology to the brain from an initial site in sympathetic nerves and ganglia. Seemingly the only anatomical pathway for this would be through the spinal cord but several groups have reported never seeing synuclein pathology in the spinal cord when it was not seen in the brain (11, 59–61). Del Tredici and Braak (59) pointedly stated, “Because the Parkinson’s disease-related lesions were observable in the spinal cord only after Lewy pathology was seen in the brain, it could be concluded that, within the central nervous system, sporadic Parkinson’s disease does not begin in the spinal cord”.

Yet, as Borghammer and colleagues point out, if there is a GI-first subtype, or if GI and brain synucleinopathy occur more or less in parallel, and if these subjects are more than a rare occurrence, it should still be possible to find, in a large autopsy series, at least a few cases with isolated GI α-synuclein pathology. The discovery of such cases may have been hampered in studies to date due to relatively sparse sampling of the GI tract. Bilateral asymmetry of CNS and PNS α-synuclein pathology might also complicate the issue although we have investigated this and found no asymmetry of peripheral synucleinopathy, at least for the submandibular gland (62).

Borghammer and colleagues also cite studies that have found a high prevalence of aggregated synuclein in the GI tract of younger normal subjects including children (63–64) but ordinarily the finding of an aging disease biomarker in large numbers of young normal subjects would bring into question its specificity for the disease. The relevance to PD of various types of synuclein deposits found in high percentages of young normal subjects is questionable unless one proposes that additional host or environmental factors are required to convert these into truly pathogenic forms.

Additionally, although we and others have previously used IHC methods that have been repeatedly demonstrated to be highly sensitive and specific for both CNS and PNS synuclein pathology, as found in multi-center blinded studies (65–69) and biochemical studies (70), it is possible that the initial form of peripheral synucleinopathy may differ from that commonly seen in the CNS. Alternate forms, among many possibilities, include α-synuclein oligomers as well as truncated and nitrated forms (71–76).

Recently, α-synuclein aggregates have become detectable by RT-QuIC and other “seeding amplification assays” (SAA)(77–83) and these may possibly be a required first step towards clinically-manifest disease, very likely preceding IHC-identified synuclein pathology. To date, these SAA methods have been underutilized in GI studies of PD (84–86) but may be more sensitive than IHC for detecting GI synuclein pathology. We therefore undertook a comprehensive SAA survey for GI synuclein seeds across 10 distinct GI sites in a large number of autopsied subjects that were either neuropathologically confirmed with PD or were clinically non-demented and without clinical parkinsonism during life.

## MATERIALS AND METHODS

### Human subjects

Recruitment, clinical assessment and autopsies took place at Banner Sun Health Research Institute (BSHRI), which is part of Banner Health, a non-profit healthcare provider centered in Phoenix, Arizona. BSHRI and the Mayo Clinic Arizona are the principal members of the Arizona Parkinson’s Disease Consortium. Brain necropsies and neuropathological examinations were performed on elderly subjects who had volunteered for the Arizona Study of Aging and Neurodegenerative Disorders (AZSAND) and BSHRI Brain and Body Donation Program (BBDP), a longitudinal clinicopathological study since 1997 (87). Procedures involving human subjects are done in compliance with the ethical standards of the Helsinki Accord and the appointed BSHRI Institutional Review Boards; all subjects or their legally-authorized representatives signed a written informed consent. The majority of subjects are clinically characterized with annual standardized test batteries that include movement disorders and cognitive/behavioral/neuropsychological components. Clinical assessment includes, amongst other tests, the Clinical Dementia Rating (CDR) Scale, Mini Mental State Examination (MMSE), Montreal Cognitive Assessment (MoCA), Unified Parkinson’s Disease Rating Scale (UPDRS) and the Scales for Outcomes in Parkinson’s Disease – Autonomic (SCOPA-AUT) For most subjects, the full neuropsychological and movement disorders battery that is part of AZSAND (87) was used to establish the relevant clinical history, presence or absence of parkinsonism or dementia and the degree of cognitive impairment. The diagnoses of mild cognitive impairment and dementia were decided by clinical consensus conferences reviewing the full test batteries, by participating neuropsychologists and cognitive/behavioral neurologists. Additionally, private medical records are requisitioned and reviewed for each subject and a postmortem questionnaire is conducted with subject contacts to help determine the presence or absence of dementia and parkinsonism for those subjects that did not have a recent standardized antemortem evaluation. All subjects in this study received whole-body autopsies with microscopic diagnoses by board-certified anatomical pathologists.

Case selection was done by searching the AZSAND database for all those that had died and had a full clinical evaluation and autopsy including 10 GI regions: submandibular gland, upper esophagus, lower esophagus, stomach, duodenum, jejunum, ileum, transverse colon, sigmoid colon and rectum. Subjects with destructive GI pathologies, including malignant neoplasms and bowel infarction or ischemia, were excluded. Subjects were chosen to have one of three clinicopathological diagnoses, PD, incidental Lewy body disease (ILBD; brain IHC-positive for synuclein pathology but clinically non-demented without parkinsonism) or controls without either dementia or parkinsonism or brain LBP (designated as No Lewy Bodies or “NLB”). Clinical diagnostic criteria for PD stipulated the presence of 2 of 3 cardinal signs (bradykinesia, rest tremor, muscular rigidity) with a beneficial response to dopaminergic replacement therapy. Postmortem diagnostic criteria for PD specified clinical parkinsonism with substantia nigra pigmented neuron loss and H & E or IHC evidence of nigral synuclein pathology.

### Histological methods

The process leading to the choice and evaluation of immunohistochemical methods for demonstrating pathological α-synuclein has been described in previous publications (65–70, 94). The standard method used at AZSAND employs proteinase K pretreatment, which not only results in superior epitope exposure but also may assist with the pathological specificity of the stain by digesting normal, non-aggregated α-synuclein, which is abundant in all nervous tissue. We use an antibody specific for α-synuclein phosphorylated at serine 129 (pSyn) (70, 89–93), which also helps identify stained structures as pathological since most clinically normal elderly subjects do not have appreciable pSyn-immunoreactive brain tissue elements, either by IHC or Western blot (70).

From each postmortem subject, one pSyn-stained slide from each of 10 brain regions was examined by a single observer (TGB) as part of the routine AZSAND diagnostic workup, including olfactory bulb, anterior medulla, two levels of pons, amygdala with adjacent entorhinal and transentorhinal areas, cingulate gyrus, inferior temporal gyrus, middle frontal gyrus and inferior parietal lobule. The presence of substantia nigra LBP was assessed in sections stained with H & E and thioflavin S, or with pSyn IHC. All cases were staged according to their brain LBP distribution with the Unified Staging System for Lewy Body Disorders (USSLB) (95) and the LBP density in each section was semi-quantitatively graded between 0 and 4 (95), with a total possible brain LBP summary score of 40.

### Preparation of gastrointestinal and brain samples

Gastrointestinal and brain samples were dissected fresh at autopsy, placed on sheets of dry ice and then stored in freezers at –70-80 C. Frozen samples were shipped on dry ice to the Rocky Mountain Laboratory in Hamilton, Montana and kept in freezers at –70 C until used. Samples were washed at least twice with 0.5 mL of TBS; occasionally an additional 1-2 washes were necessary to reduce, for example, blood or bile contamination. The tissues were weighed, placed in a tube with 3 x 3 mm borosilicate glass beads (Chemglass Life Sciences, Germany; Cat# CG-1101-02) and TBS containing 2 mM CaCl_2_ with 0.25% (w/v) collagenase A (final 10% w/v concentration). Tissues were minced for 1 min at maximum speed in a Mini-Beadbeater. Then samples were incubated for 2 h at 37°C with 400 rpm shaking. Next, a second 1-min maximum speed round of homogenization with the Mini-Beadbeater was performed. The homogenates were then centrifuged for 2 min at 2,000x g and the supernatant was dispensed in single use aliquots to be stored at –70°C.

Brain samples were homogenized at 10% (w/v) in a tube with 1 mm zirconia beads and PBS, using a Mini-Beadbeater (1 min at maximum speed), then centrifuged for 2 min at 2,000 x g. The supernatant was distributed in single use aliquots and stored at –70°C.

### Alpha-synuclein seed amplification assay (SAA) methods

Clear-bottom 96-well plates (Thermo Fisher Scientific, Waltham, MA, USA; Cat# 1256672) were preloaded with six 800-µm silica beads (OPS Diagnostics, Lebanon, NJ, USA; Cat# 80020002) per well. The reaction mixture was prepared fresh with 0.1 mg/mL (measured via NanoDrop) of in-house purified mutant αSyn K23Q-αSyn monomer (Accession No. NM_000345.3) filtered with a 50 kDa Amicon filter (Millipore Cat# UFC5050BK), 10 µM thioflavin T (ThT), 40 mM phosphate buffer (pH 8.0), and 170 mM NaCl. Reaction mixture (98 µL) was dispensed into each well, followed by 2 µL of a 1:100 dilution of 10% GI or brain tissue homogenates. Samples were tested in ≥ 4 replicate reactions and plates were sealed with a plate sealer (Thermo Fisher Scientific, Waltham, MA, USA; Cat# 235307). Reactions were incubated in a BMG FLUOstar Omega (BMG Labtech, Ortenberg, Germany) reader at 42°C (1 min 400 rpm double orbital shaking followed by 1-min rest) with ThT fluorescence readings (445 nm excitation and 480 nm emission wavelengths), every 45 min.

The primary criterion used to label a sample positive was the number of replicate reactions crossing the designated fluorescence threshold, defined as 10% of the maximum value on each plate within an assay incubation time of 30 hours. Samples were tested in quadruplicate and classified as 4/4, 3/4, 2/4, 1/4, or 0/4 positive replicates; when an SAA-tested sample was positive in 4/4 or 3/4, it was defined as overall positive without retesting. Samples scoring 2/4 were retested, whereafter retested samples scoring ≥ 2/4 were defined as positive; otherwise, samples scoring 1/4 or 0/4 were considered negative.

### Hypotheses and statistical tests

We sought to test the hypothesis that LBP in the GI tract, whether detected by pSyn IHC or SAA, may at least occasionally occur in the absence of brain LBP, and to determine, if any cases of “GI-only” LBP are detected, the prevalence of these as compared with “Brain-Only” cases. A stepwise process was devised for this at the study planning stage (Figure 1). Group proportions in the three diagnostic groups were compared with 2-way Fisher exact tests. Group means were compared with analysis of variance (ANOVA) and post-hoc Tukey’s tests or Kruskal-Wallis analysis of variance and post-hoc Dunn’s tests.

**Figure 1.**
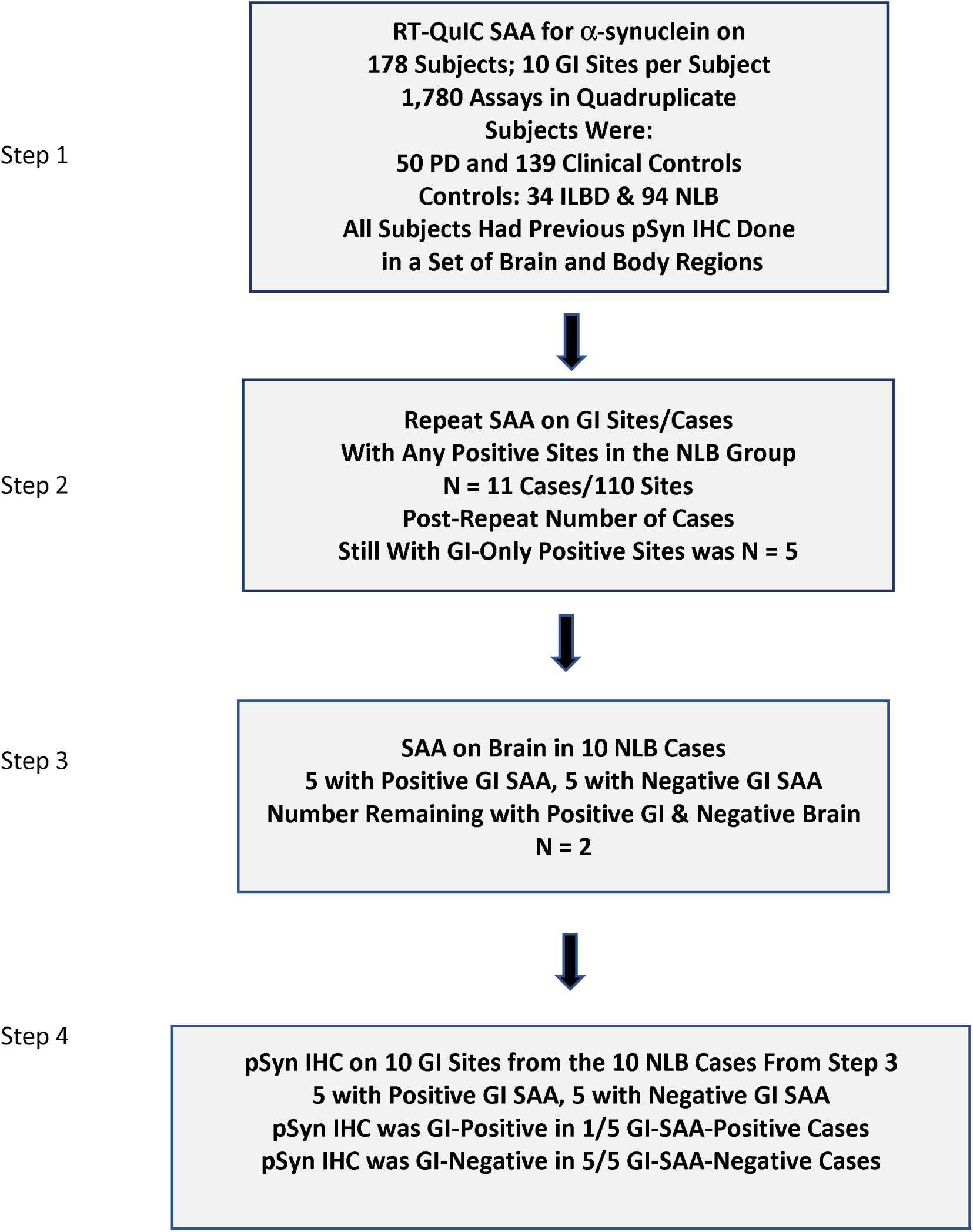
Experimental plan. A stepwise process was implemented. SAA = Alpha-synuclein seed amplification assay; PSyn = phosphorylated alpha-synuclein; IHC = immunohistochemistry.

## RESULTS

### Subject characterization – general and parkinsonism-related

Subjects included 50 clinicopathologically diagnosed with PD, 34 with ILBD and 94 with NLB (Table 1; Supplementary File). The subjects were predominantly of advanced age, with the mean age ranges for the diagnostic groups falling between 80 and 87 years. Median disease duration for PD cases was 12 years, ranging from 2 to 44 years. PD cases were significantly younger than NLB or ILBD cases, were more likely to be male when compared to the normal group and had significantly higher UPDRS motor and total SCOPA-AUT scores than the other groups. The median postmortem intervals ranged between 3.0 and 3.5 hours with no significant differences amongst groups.

**Table 1.** Clinical characteristics of study subjects, by clinicopathological diagnosis, age, sex, last motor UPDRS score and disease duration. Means and standard deviations (SD) are given except for PMI (medians). ILBD = incidental Lewy body disease; PD = Parkinson’s disease; NLB = non-demented and without parkinsonism with no Lewy body pathology; PMI = postmortem interval in hours; UPDRS = last Unified Parkinson’s Disease Rating Scale, Part III total score; SCOPA = last SCOPA-Autonomic total score; Dis Dur = disease duration in years. * PD cases were significantly younger than Normal (p < 0.001) or ILBD cases (p = 0.0009), were more likely to be male when compared to the Normal group (p < 0.001) and had higher UPDRS and SCOPA scores than both other groups (p < 0.0001 for both score types). The groups were not significantly different in PMI.

| **Diagnosis (N)** | **Age at Death\*** | **Sex M/F\* (%M)** | **PMI** | **UPDRS\*<sup>1,2</sup>** | **SCOPA\*<sup>3</sup>** | **Disease Duration** |
| --- | --- | --- | --- | --- | --- | --- |
| NLB (94) | 86.2 (9.2) | 38/56 (40%) | 3.1 | 13.7 (12.8) | 14.3 (7.4) | N/A |
| ILBD (34) | 87.0 (7.4) | 20/14 (58%) | 3.0 | 5.6 (5.3)<sup>2</sup> | 11.9 (6.2) | N/A |
| PD (50) | 80.1 (7.3) | 35/15 (70%) | 3.5 | 40.6 (16.1)<sup>3</sup> | 19.9 (7.25) | 13.8 (9.9) |

1. For 23 normal and 3 ILBD cases, UPDRS scores were not available. 2. For all subjects, UPDRS score shown is off medication (n = 36). 3. For 26 Normal, 6 ILBD and 15 PD cases, SCOPA scores were not available.

### Subject characterization – gastrointestinal symptoms

The subset of the SCOPA-AUT questionnaire that is focused on GI-related symptoms was compared across the three diagnostic groups (Figure 2). Single category scores, ranging between 0 and 3 for each category, had significant group differences in those relating to excess salivation, straining at stool, constipation and decreased frequency of bowel movements. For three out of four of these categories, scores for the PD group were significantly different from both the NLB and ILBD groups. The bowel movements score was significantly different between PD and NLB but not between PD and ILBD. Differences between NLB and ILBD were not statistically significant for any of the four categories.

**Figure 2.**
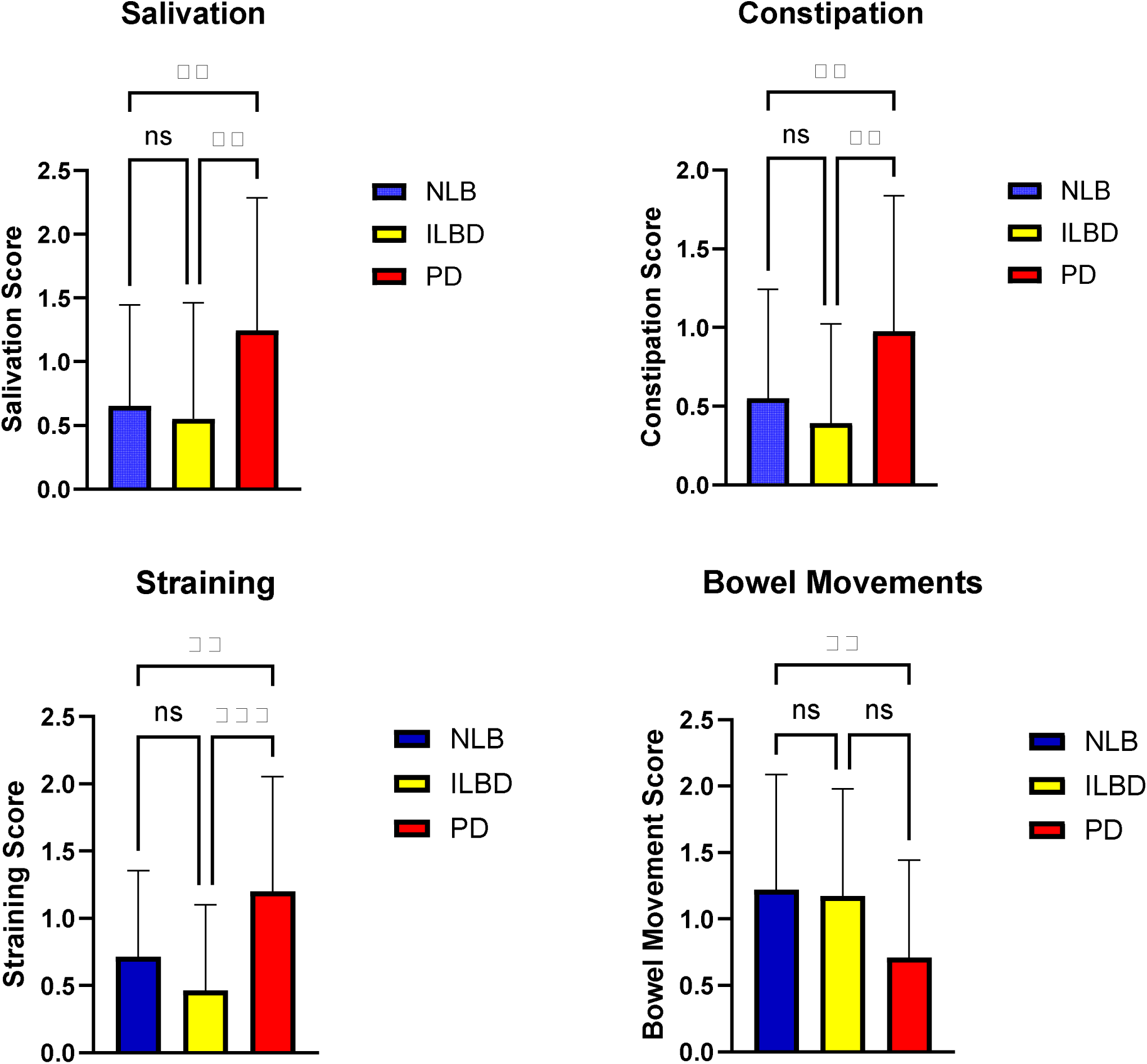
Comparison between diagnostic groups of GI-related results on the SCOPA-AUT questionnaire. Significant single category score differences (Kruskal-Wallis with post-hoc pairwise Dunn’s tests) were present for 4 gastrointestinal symptoms: those relating to excess salivation, straining at stool, constipation and decreased frequency of bowel movements. ** p < 0.01, *** p < 0.001.

### Step 1 & Step 2: GI SAA-positive NLB subjects and sites

Step 1 of the SAA regional analysis (Figure 1) initially resulted in 11 GI SAA-positive NLB subjects, thus potentially 11 GI-only LBD cases. After repeat SAA of all samples from these cases (Figure 1, Step 2), 6 cases were found consistently negative, giving a remaining total of 5 GI SAA-positive NLB subjects, and thus potentially 5 GI-only LBD cases. Most of these subjects were positive at only one or two sites but one subject was positive in 6/10 sites.

### Step 3: Results of brain α-synuclein SAA on GI-only SAA-positive NLB subjects

As the brains from the 5 GI SAA-positive NLB subjects had only been assessed by IHC for synucleinopathy, and IHC is likely to be less sensitive than SAA, it was uncertain whether or not these 5 cases were truly “GI-only” with respect to their synucleinopathy. Step 3 (Figure 1) of the project therefore performed α-synuclein SAA on four brain regions on 10 subjects from Step 2 including the 5 GI-positive subjects as well as 5 GI-negative subjects. The chosen brain regions were selected for their high frequency of synucleinopathy (95) in Lewy body disorders, including olfactory bulb/tract, anterior medulla with dorsal motor nucleus of the vagus nerve, pons with locus ceruleus and medial amygdala at the uncus. The results found amygdala to be SAA-positive in 3 subjects, medulla in 2, and olfactory bulb/tract in 1. Collectively, three subjects that had apparent GI-only positive SAA results had positive brain SAA results, leaving only 2 subjects with GI-only SAA positivity. One of these, a woman in her 80s, was positive in only one GI site, the rectum, while the other, a man in his 80s, was positive in 6 sites: submandibular gland, upper and lower esophagus, stomach, jejunum and rectum. Both were judged to be normal for age proximal to death, in terms of both cognition and movement.

### Results of GI SAA positivity rates in diagnostic groups and GI sites

Initially (Figure 1, Step 1), of 1,760 assays of 10 GI sites of all 178 subjects, 619 sites were considered positive. Of the three diagnostic groups, PD subjects had the highest proportion of positive subjects (48/50; 96%) and GI sites (453/500; 90%), followed by ILBD subjects (24/34; 71%) and sites (130/350; 37%) and NLB subjects (11/94; 11%) and sites (23/940; 2%); the proportions are significantly different (p < 0.0001, Fisher’s exact test) and the diagnostic groups also significantly differ in the numbers of positive sites per subject (Figure 3). The regional variation in positivity rates, best seen with the combined NLB and ILBD groups (Figure 4), have a rostrocaudal gradient, being greatest (p = 0.0048, Fisher’s exact test) in the upper digestive tract regions (submandibular gland, esophagus, stomach) as compared to the lower regions (duodenum, ileum, jejunum, colon and rectum).

**Figure 3.**
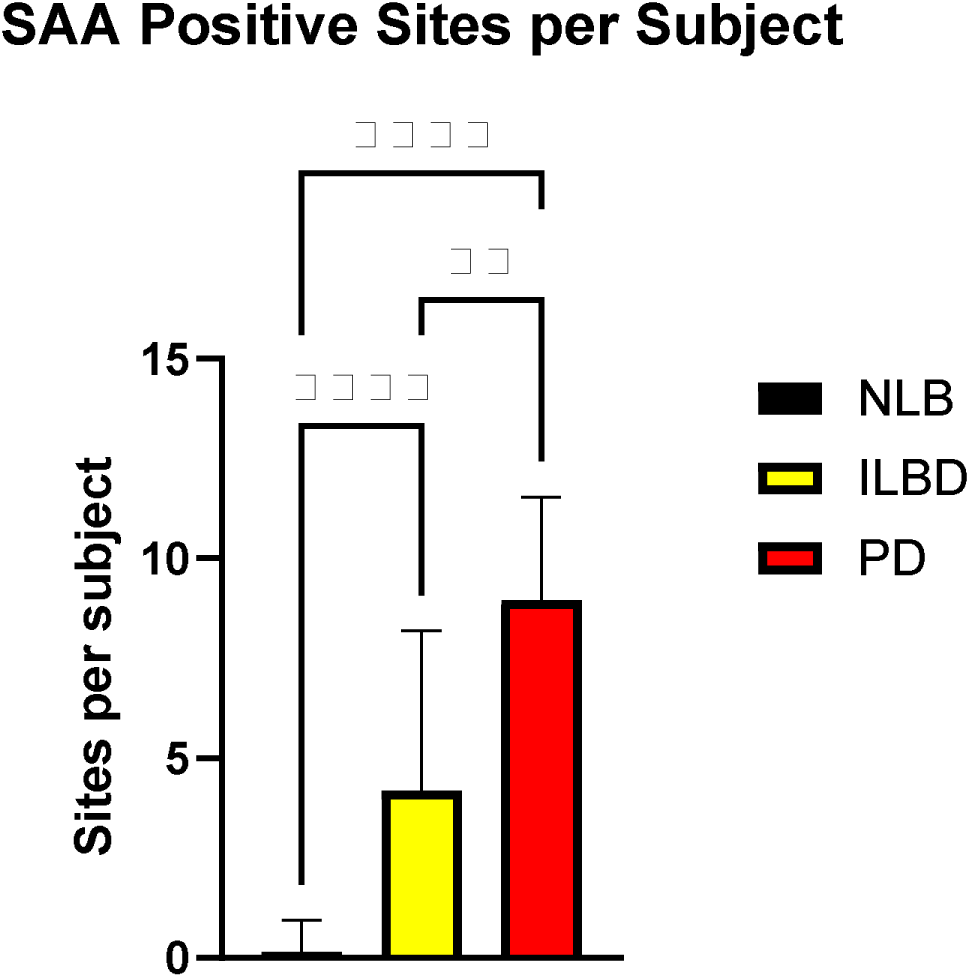
The mean number of positive α-synuclein seeding amplification assay (SAA) GI sites is significantly different between the three diagnostic groups (Kruskal-Wallis with post-hoc pairwise Dunn’s tests). ** p < 0.01, **** p < 0.0001.

**Figure 4.**
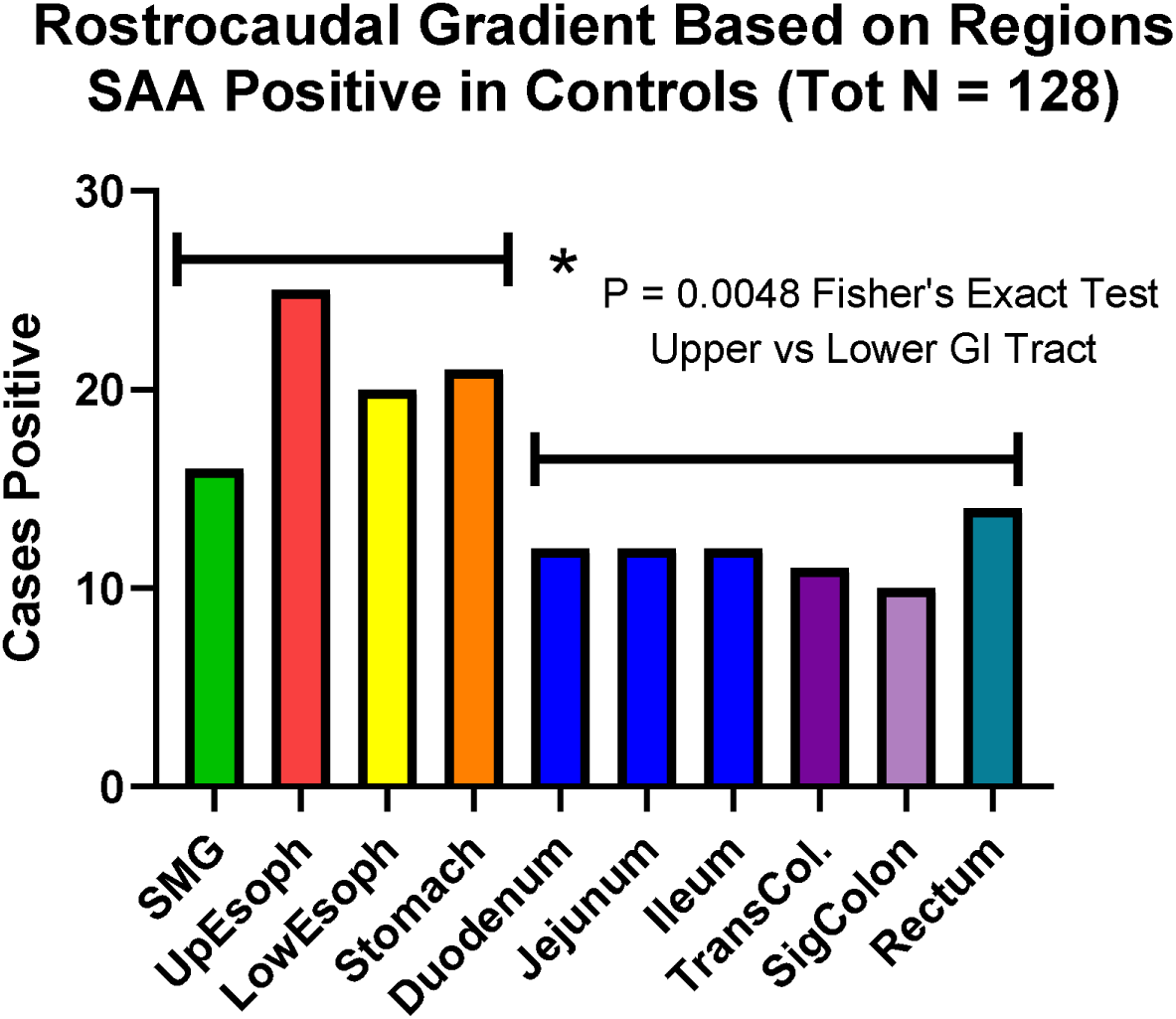
The regional GI positivity rate with α-synuclein seeding amplification assays, in the combined non-Lewy pathology Control group and ILBD groups, roughly followed a rostro-caudal gradient. The mean positivity rate in the upper GI tract regions is significantly greater than the mean positivity rate in the lower GI tract regions (p = 0.0048, Fisher’s exact test).

### Correlations of GI SAA positivity with brain pSyn IHC burden & clinical and autopsy conditions

As each subject has SAA results for 10 GI regions, the number of positive regions in each subject was used as a semi-quantitative measure of the total GI synuclein aggregate burden. Figure 5 shows that the number of SAA-positive GI regions per subject is significantly associated with the total pSyn brain LBP regional summary score (Spearman rho = 0.89, p < 0.0001) as well as with the UPDRS motor score (Spearman rho = 0.44, p < 0.0001).

**Figure 5.**
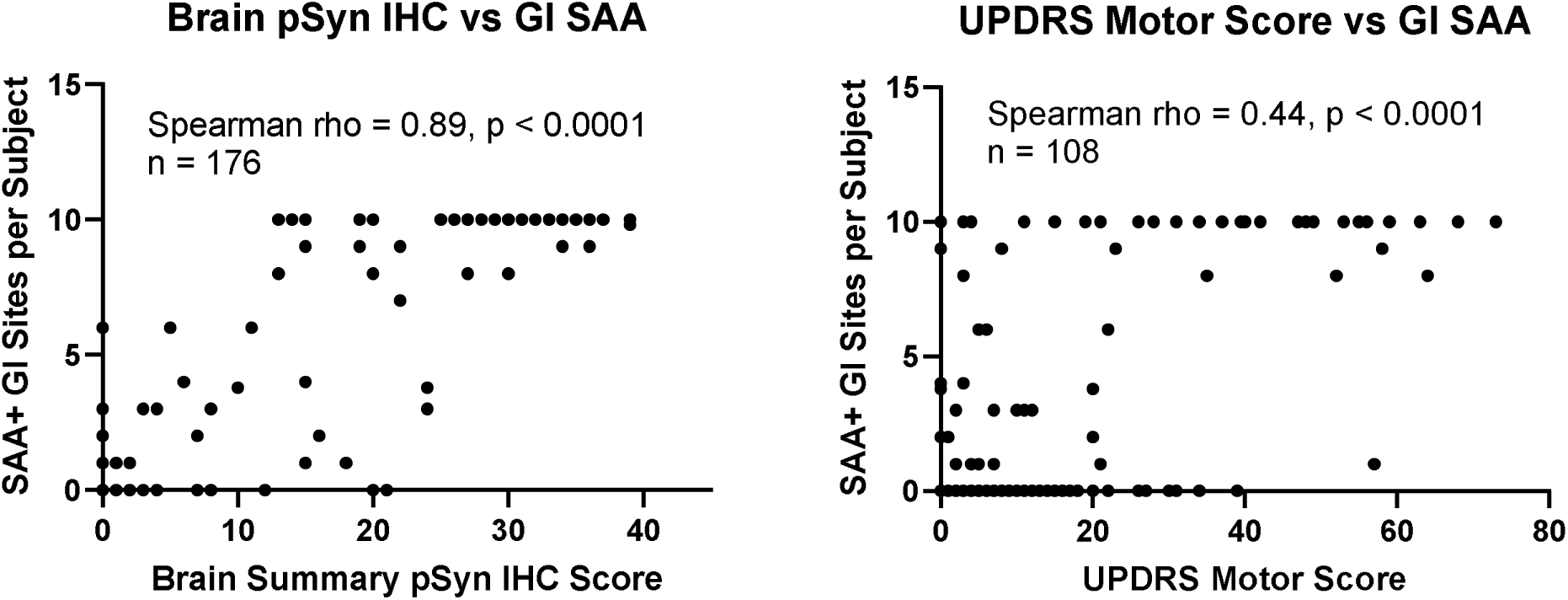
The number of SAA-positive GI regions per subject, including all subjects, is significantly associated (Spearman rho = 0.89, p < 0.0001) with the total pSyn IHC brain regional summary score, as well as with the UPDRS motor score (Spearman rho = 0.44, p < 0.0001).

Figure 6 shows that the number of SAA-positive GI regions per subject is significantly correlated with individual GI symptom scores included in the SCOPA-AUT questionnaire, including bowel movements (Spearman rho = – 0.32, p < 0.0001), constipation (Spearman rho = 0.19, p = 0.02), and salivation (Spearman rho = 0.17, p = 0.034). The correlation with total SCOPA-AUT score is not significant, however.

**Figure 6.**
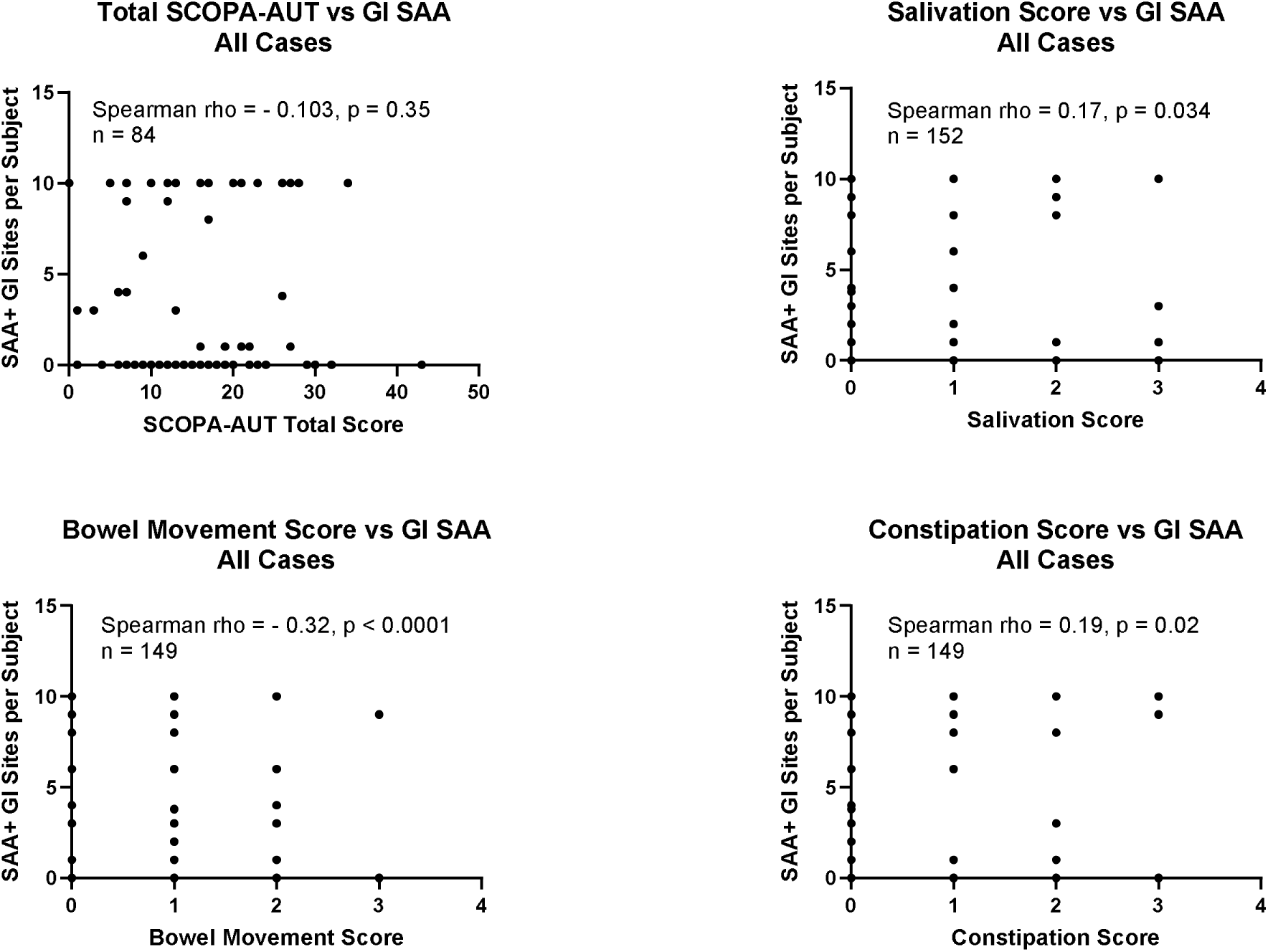
The number of SAA-positive GI regions per subject is significantly correlated with individual GI-related questionnaire items, including bowel movements (Spearman rho = – 0.32, p < 0.0001), constipation (Spearman rho = 0.19, p = 0.02) and salivation (Spearman rho = 0.17, p = 0.034), but not with the total SCOPA-AUT score.

### Step 4: Results of pSyn IHC on 10 GI sites from 10 normal Control subjects from Steps 2 & 3

Additional slides were newly stained and examined for pSyn immunoreactivity from 10 GI sites of each of 10 NLB subjects identified after Step 1 (Figure 1) that were found to be GI-only SAA-positive in one or more GI sites (after repeat GI SAA, in Step 2, only 5 were determined to be GI-only SAA-positive, and after brain region SAA of these, only two of these were ultimately found to be GI-only SAA-positive). From each of these 10 subjects, pSyn staining and examination was done on 4-6 slides from each of the same 10 GI regions used for SAA, for a total of 440 additional GI pSyn-stained slides (Figure 1, Step 4). As in our previous investigations (11, 55, 67–69), only pSyn IHC staining that was morphologically consistent with nerve fibers or ganglion cells was considered to be specific for GI synuclein pathology. Of these newly-stained slides, only two slides were positive, both within myenteric plexus of lower esophagus in a single NLB case that was SAA-positive in multiple GI sites as well as in amygdala and medulla.

It is evident that GI SAA is more sensitive for GI synucleinopathy than pSyn. This is best demonstrated by the results for ILBD subjects, for whom 71% (24/34) were GI SAA positive vs only 41% (14/34) that were GI IHC-positive. Immunohistochemistry therefore seems about 30% less sensitive than SAA, although the IHC assessment is somewhat disadvantaged in this comparison as it only assessed 2 GI regions (submandibular gland and lower esophagus) in most cases (an additional 15 cases were also assessed in stomach) while 10 GI sites were assessed by SAA.

## DISCUSSION

Braak and others proposed that PD may start from an initial site in the GI tract, with subsequent passage to the central nervous system (CNS) through the vagus nerve or other routes. We initially tested this hypothesis by using pSyn IHC in two large studies (11,55), finding no evidence for pSyn Lewy pathology selectively localized to the GI tract. Borghammer and colleagues later (50–54) proposed the existence of two subtypes of PD, one that is “brain-first” and one that is “body-first”. However, their presented evidence for this is entirely based on theoretical models, with the only empirical, autopsy-based support coming from a few Japanese studies (56–58) finding isolated pSyn IHC synuclein pathology in sympathetic nerves or ganglia of 9/178 elderly subjects (56), but still with no GI-only cases. Spread from the sympathetic nervous system to the CNS would seemingly require initial passage through the spinal cord but several groups, including Del Trecici and Braak, have reported never seeing synuclein pathology in the spinal cord when not also seen in the brain (11, 59–61).

Still, it has been pointed out that the discovery of GI-only pathology cases may have been hampered in studies to date due to relatively sparse sampling of the GI tract. We therefore undertook a comprehensive survey, using synuclein SAA, to search for GI-only synuclein seeds across 1,780 GI sites in 178 autopsied subjects that were either neuropathologically confirmed with PD or ILBD, or were clinically non-demented and without clinical parkinsonism or autopsy evidence of CNS synuclein pathology. We used synuclein SAA for this survey as it is currently thought to be the most sensitive method to detect even the very earliest signs of synuclein pathology.

The results showed that synuclein aggregates were present in the overwhelming majority of GI sites in subjects with clinicopathologically diagnosed PD, with 48/50 cases positive, in a total of 453/500 GI sites. Incidental Lewy body disease subjects were positive in 24/34 cases, in a total of 130/359 sites.

Subjects with no apparent CNS Lewy pathology, when assessed with the standard BBDP pSyn IHC diagnostic panel, termed NLB here, were SAA-positive in 11/94 subjects and 23/940 GI sites. However, after repeat GI SAA as well as brain SAA, only two subjects were confirmed as GI-only. Of the same 94 NLB subjects, synuclein pathology, by either SAA or IHC, was restricted to the brain in 11 cases. Further SAA of brain regions in NLB cases is therefore estimated, based on the proportional results to date, to potentially identify 21 additional brain-only LBP subjects (total of 32). From this, brain-only LBP is tentatively estimated to be 16 times more common (32/2) than GI-only LBP, but an accurate estimate of this fraction can only be obtained after brain SAA is done on all NLB cases in this series.

Uncertainty still exists, however, about the relevance of a positive SAA tissue result. Our previous experience has been that peripheral pSyn IHC is only specific for Lewy body CNS disease when it is clearly localized to neuronal structures (67). Unlike in the brain, where it is restricted to neurons, we have found that GI pSyn may be detected in non-neuronal cell types (96), and others have reported that pSyn and even synuclein aggregates may be ubiquitous in the normal GI tract and/or be induced there by inflammation (15, 63,64,97). A positive GI SAA result does not inform us of its cellular location and so its relationship to PD is not necessarily clear. Would synuclein aggregates always proceed to pSyn IHC or even H & E apparent Lewy bodies? At present this is unknown.

We found that the number of SAA-positive GI sites per subject is significantly correlated with the UPDRS motor score and SCOPA-AUT GI-related scores for salivation, straining, constipation and bowel movement frequency but our sample size is insufficient to determine if this correlation would be independently significant if CNS pSyn load was included in the equation. It is reasonable to assume that synuclein pathology in the enteric nervous system would eventually result in loss of its constituents and motility but we have previously found there to be no loss of enteric neurons in PD (25).

In conclusion, we have found little empirical, autopsy-based support for the theoretical concept of two types of PD, “brain-first” and “body-first”. Instead, it seems likely that, in the great majority of people with PD, synuclein pathology begins in the brain, and only “spreads” to the GI tract after reaching a threshold of regional positivity in the brain. The existence of a very small percentage of subjects with isolated PNS sympathetic neuronal synuclein pathology is not surprising, considering the selective vulnerability of CNS catecholaminergic neurons to synuclein pathology, but it is unclear how synuclein pathology in peripheral sympathetic neural elements would ultimately be transmitted to the brain. Such isolated PNS sympathetic neuronal synucleinopathy might represent preclinical cases of pure autonomic failure rather than preclinical PD.

## FUNDING

This study was funded by a grant from the Michael J. Fox Foundation for Parkinson’s Research (MJFF-022763). The Arizona Study of Aging and Neurodegenerative Disorders and Brain and Body Donation Program has been supported by the National Institute of Neurological Disorders and Stroke (U24 NS072026 National Brain and Tissue Resource for Parkinson’s Disease and Related Disorders), the National Institute on Aging (P30 AG019610 and P30AG072980, Arizona Alzheimer’s Disease Center), the Arizona Department of Health Services (contract 211002, Arizona Alzheimer’s Research Center), and the Arizona Biomedical Research Commission (contracts 4001, 0011, 05-901 and 1001 to the Arizona Parkinson’s Disease Consortium). This work is the result of NIH funding, in whole or in part, and is subject to the NIH Public Access Policy. Through acceptance of this federal funding, the NIH has been given a right to make the work publicly available in PubMed Central. This research was also supported by the Intramural Research Program of the National Institutes of Health (NIH). The contributions of the NIH authors were made as part of their official duties as NIH federal employees, are in compliance with agency policy requirements, and are considered Works of the United States Government. However, the findings and conclusions presented in this paper are those of the authors and do not necessarily reflect the views of the NIH or the U.S. Department of Health and Human Services. The funding body played no role in the design of the study and collection, analysis, and interpretation of data and in writing the manuscript.

## CONFLICTS OF INTEREST

BC and CDO report ownership of patents related to a-synuclein SAA technologies. All of the other authors report no direct financial conflicts of interest related to this project.

## Data Availability

All data produced in the present work are contained in the manuscript and supplemental files.

## ACKNOWLEDGEMENTS

We thank Lori Lubke for her technical assistance and diligent record keeping. We are grateful to all of the research subjects who volunteered their time and commitment, as well as their bodies, to this project.

We could not have completed this project without the many dedicated and skilled clinical and technical staff that have served the Arizona Study of Aging and Neurodegenerative Disorders over the past 29 years.

## AUTHOR ROLES

Charles H. Adler: Design, execution, analysis, writing, editing of final version of the manuscript.

Parvez Alam: Execution, editing of final version of the manuscript.

Alireza Atri MD, PhD: Execution, editing of final version of the manuscript.

Thomas G. Beach: Funding, design, execution, analysis, writing, editing of final version of the manuscript.

Christine M. Belden: Execution, editing of final version of the manuscript.

Byron Caughey: Execution, analysis, writing, editing of final version of the manuscript.

Erika Driver-Dunckley: Execution, editing of final version of the manuscript.

Bradley Groveman: Execution, editing of final version of the manuscript.

Andrew G. Hughson: Execution, analysis, editing of final version of the manuscript.

Anthony Intorcia: Execution, editing of final version of the manuscript.

Samantha King: Execution, editing of final version of the manuscript.

Ileana Lorenzini: Execution, editing of final version of the manuscript.

Sabiha Parveen: Execution, editing of final version of the manuscript.

Sanaria H. Qij: Execution, editing of final version of the manuscript.

Holly A. Shill: Execution, editing of final version of the manuscript.

Shyamal Mehta: Execution, editing of final version of the manuscript.

Christina D. Orrú: Execution, analysis, writing, editing of final version of the manuscript.

Geidy E. Serrano: Design, execution, analysis, writing, editing of final version of the manuscript.

Sarah Vascellari: Execution, editing of final version of the manuscript.

## Notes

### Competing Interest Statement

BC and CDO report ownership of patents related to alpha synuclein SAA technologies. All of the other authors report no direct financial conflicts of interest related to this project.

### Funding Statement

This study was funded by a grant from the Michael J. Fox Foundation for Parkinson Research. The Arizona Study of Aging and Neurodegenerative Disorders and Brain and Body Donation Program has been supported by the National Institute of Neurological Disorders and Stroke U24 NS072026 National Brain and Tissue Resource for Parkinson Disease and Related Disorders the National Institute on Aging P30 AG019610 and P30AG072980 Arizona Alzheimer Disease Center the Arizona Department of Health Services contract 211002 Arizona Alzheimer Research Center and the Arizona Biomedical Research Commission contracts 4001 0011 05-901 and 1001 to the Arizona Parkinson Disease Consortium. This work is the result of NIH funding in whole or in part and is subject to the NIH Public Access Policy. Through acceptance of this federal funding the NIH has been given a right to make the work publicly available in PubMed Central. This research was also supported by the Intramural Research Program of the National Institutes of Health NIH. The contributions of the NIH authors were made as part of their official duties as NIH federal employees are in compliance with agency policy requirements and are considered Works of the United States Government. However the findings and conclusions presented in this paper are those of the authors and do not necessarily reflect the views of the NIH or the U.S. Department of Health and Human Services. The funding body played no role in the design of the study and collection analysis and interpretation of data and in writing the manuscript.

### Author Declarations

The Western Institutional Review Board of Puyallup WA gave ethical approval for this work Study Number 1132516 Protocol Number 20120821.

